# Future scenarios for the SARS-CoV-2 epidemic in Switzerland: an age-structured model

**DOI:** 10.1101/2020.05.28.20115659

**Authors:** Janne Estill, Plamenna Venkova-Marchevska, Maroussia Roelens, Erol Orel, Alexander Temerev, Antoine Flahault, Olivia Keiser

## Abstract

The recent lifting of COVID-19 related restrictions in Switzerland causes uncertainty about the future of the epidemic. We developed a compartmental model for SARS-CoV-2 transmission in Switzerland and projected the course of the epidemic until the end of year 2020 under various scenarios. The model was age-structured with three categories, children (0-17), adults (18-64) and seniors (65- years). Lifting all restrictions according to the plans disclosed by the Swiss federal authorities by mid-May resulted in a rapid rebound in the epidemic, with the peak expected in July. Measures equivalent to at least 90% reduction in all contacts were able to eradicate the epidemic; 56% reduction in contacts could keep the intensive care unit occupancy under the critical level, and delay the next wave until October. Scenarios where strong contact reductions were only applied in selected age groups could not suppress the epidemic, increasing the risk of a next wave in July, and another stronger wave in September. Future interventions need to cover all age groups to keep the SARS-CoV-2 epidemic under control.

---

Switzerland has one of the highest incidences of documented SARS-CoV-2 infections per population, with large regional variability.[1,2] In response to the SARS-CoV-2 pandemic, the Swiss federal government implemented several restrictions, including closures of schools and non-essential shops and services and forbidding gatherings of more than five people.[3] Many of these restrictions were lifted on May 11, 2020, causing uncertainty about the future of the epidemic. We have developed an age-structured mathematical model to estimate possible scenarios for Switzerland until December 2020, and to identify how different levels of contact reduction between different age groups influence SARS-CoV-2 transmission.

We used a stochastic compartmental model (code and a detailed description of the methodology are available in the Appendix). The population is divided in three age groups; children (0–17 years), adults (18–64 years) and seniors (≥65 years). The model was parameterised using literature estimates from a model for France, and fitted against the daily COVID-19 hospitalizations and deaths.[2,4] Before running the model for Switzerland, we fitted it to three cantons with epidemics that started at different times (Geneva, Ticino and Bern) to select informative priors for uncertain parameters. We included the impact of the government-imposed restrictions. We modelled seven scenarios. We assumed in all scenarios that strong social distancing (restricting all larger gatherings of adults) would continue until 7 June, and “light” social distancing (awareness, hand hygiene) for the rest of the year. Relative reductions in contacts were calculated from this baseline assumption. Contacts are defined as all contact scenarios in which transmission is possible from a single infectious person, regardless of type or duration. We represented future interventions (including contact tracing, intensified screening, and wearing masks) by an overall reduction in either all contacts or contacts between specific age groups. We present the means of 1000 simulations with 95% credible interval (CrI).

Easing of restrictions may lead to a rapid increase in infections from late June, if the population relaxes social distancing and no effective tracing or testing efforts are implemented. In this scenario, 83% of the Swiss population would become infected by the end of the year (Figure). By restricting total contacts by 90%, we estimate a full eradication of the epidemic resulting in no infectious individuals in Switzerland by 25 July. However, low overall immunity (<5%) leaves the population vulnerable to new outbreaks. A 56% reduction in all contacts would retain the occupancy of ICU beds below the current maximum capacity (about 1200)[5] throughout the year. In this scenario, the effective reproductive number *R*_e_ would stay around 1.4, decreasing to <1 only during summer holidays. In this scenario, a new wave would start in October and only 5.2% (95% CrI 3.4–7.6%) of the population would have been infected by the end of 2020.

**Figure.**
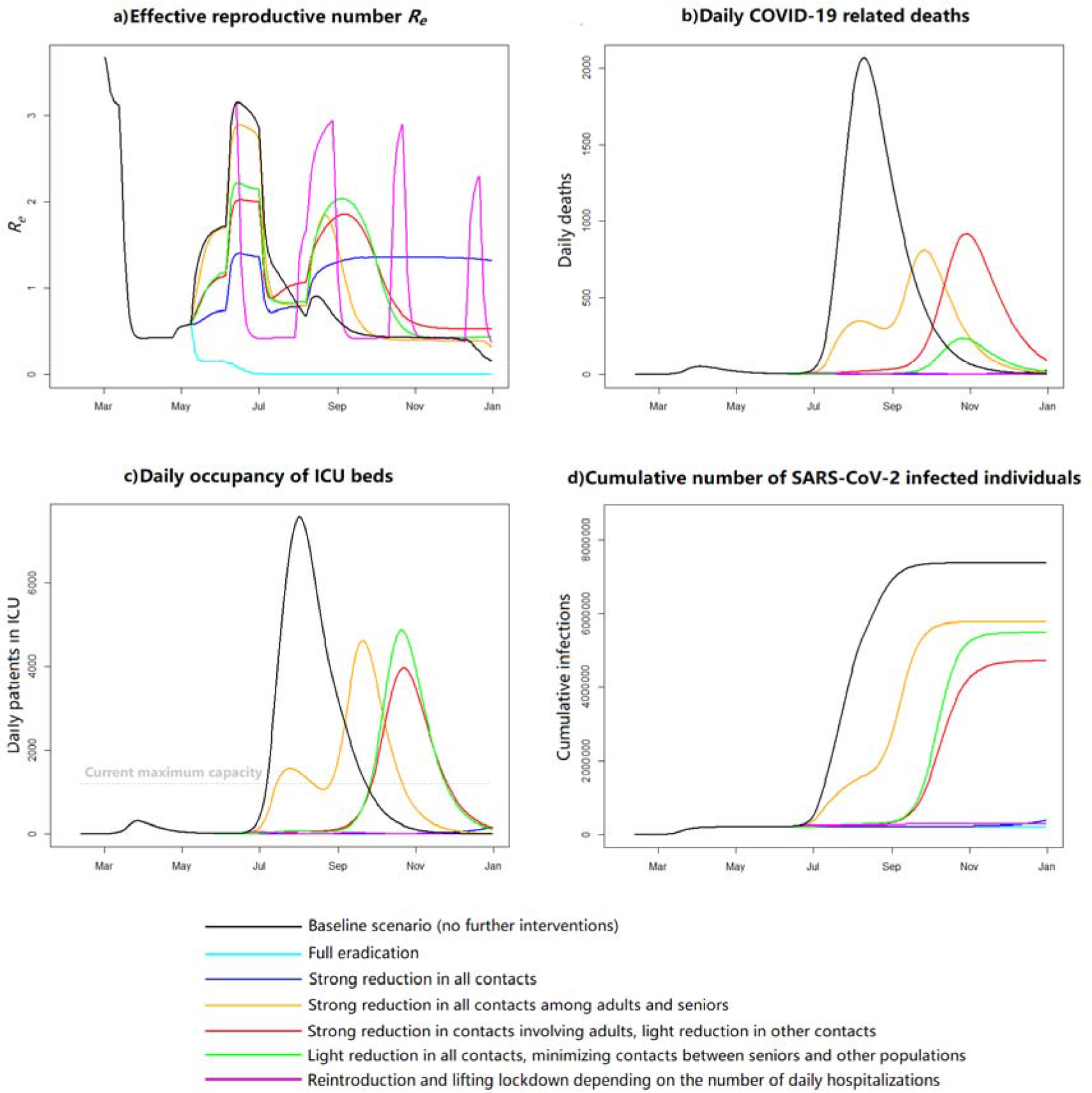
Results from different scenarios: a) Reproductive number, b) daily COVID-19 related deaths, c) daily intensive care unit (ICU) bed occupancy, and d) cumulative infections in Switzerland from 11 February to 31 December 2020. “Strong reduction” refers to a 56% reduction of contacts (calibrated to prevent ICU overflow with 90% probability), “light reduction” is half of that, and “minimizing contacts” refers to 95% reduction of contacts.

Maintaining a minimum of 56% contact reduction for all age groups is essential. Applying this reduction only for contacts among adults and seniors, without restricting contacts involving children, would result in two peaks, in late July and late September, and a substantial ICU overflow and mortality from mid-July until mid-October. In this scenario, 64.9% (95% CrI 63.6–66.2%) of the population would have been infected by the end of 2020. When reducing contacts involving adults by 56%, and contacts among children and seniors only by 28%, we predict no peak in July but a larger peak in autumn, resulting in a similar disease burden (cumulative proportion of infected individuals 54.8%, 95% CrI 54.7–54.9%). A similar pattern in the epidemic, with the next strong peak in October, was also seen when we restricted all contacts by 28%, except those between seniors and other age groups by 95%. In this case the number of deaths was however four times lower and the cumulative proportion of infected higher (63.3%, 95% CrI 63.3–63.6%). All above-mentioned scenarios were sensitive to the parameterisation: for example, in the model calibration for the Canton of Geneva, restricting contacts among adults and seniors only resulted in a stronger peak during summer without the second peak, whereas the bimodal pattern was in turn seen in the two other scenarios.

In the last modelled scenario, we assumed that all lockdown measures (as between 20 March–26 April) would be reintroduced immediately if the number of daily new hospitalizations grew to 40, and lifted two weeks after they returned back below 10. This criteria would result in four new lockdowns: 16 June–1 August 2020, 31 August–14 October 2020, 25 October–13 December 2020, and 24 December 2020–13 February 2021. The number of patients in ICU remained below 200 throughout. Overall 5.6% (95% CrI 4.8–6.6%) of the total population would have been infected by 31 December.

We also conducted a number of sensitivity analyses. The scenarios were sensitive to the duration of the latency period and infectiousness. The shorter the serial interval, the easier it will be to eradicate the epidemic or delay the second wave. Seasonal forcing, which is a potential but currently disputed factor,[6,7] could also slow down the epidemic: assuming a strong seasonality factor could delay the next wave to at least September.

Our study demonstrates that as long as the virus is present in a community with limited immunity, there is a risk of a rapid rebound of the epidemic if the restrictions are lifted and the people stop following social distancing and other protective behaviour. In the absence of seasonal forcing, the next wave could in theory occur in the summer. Efforts to control the epidemic, such as intensive testing, contact tracing, wearing of masks and hygiene measures, must have sufficient coverage to limit transmission among all age groups. No single intervention is enough. Reducing transmission uniformly may keep the epidemic suppressed only until late autumn. Restricting contacts between seniors and the rest of the population will prevent deaths, but is not sufficient to control the epidemic. A “start-stop” strategy where the trends in COVID-19 related hospitalizations (or confirmed cases) trigger a new lockdown, is effective but not feasible: this would push the lockdown into the future, with only a few weeks of “normality” in between.

This study provides a broad range of future scenarios. In reality, the most severe scenarios are very unlikely: we can expect that the public health authorities will react and the people will adapt their behaviour if there is a new increase in cases. Our study has also several limitations. We focused on the average restriction of transmission-supporting contacts, without considering the practical implementation of specific interventions. The baseline contact patterns were based on literature data. They may however not fully reflect transmission patterns as the knowledge on transmission routes is still limited. Some contacts may be easier to track and control than others. Using a compartmental model, we could not differentiate between household and community transmission, or consider the effect of superspreaders.

An effective response to the COVID-19 pandemic needs several components: the spread of the infection must be kept as low as possible, the vulnerable population groups need additional protection, effective monitoring strategies must be in place, and the society must be ready to reintroduce additional measures if necessary. Only a combined prevention approach that targets all population groups can assure a sufficient control of the epidemic until a vaccine becomes available.

## Data Availability

The data used for this project are collected from publicly available sources (see description in the manuscrpit, or the repository https://gitlab.com/igh-idmm-public/covid-19/modelling_jestill)

## Acknowledgments

We thank Rachel Esra, University of Geneva, for editing the paper.

